# Longitudinal multi-omics characterization of the malignant evolution in multirelapsing glioblastoma

**DOI:** 10.64898/2026.06.11.26355035

**Authors:** Madeleine H. Lackman, Christopher Wardell, Emilie Darrigues, Annick De Loose, A. Geoffrey Lyle, Yuanqing Xue, Katrina Learned, Allison Cheney, Olena M. Vaske, Sinem Karaman, Vadim Le Joncour, Analiz Rodriguez

**Affiliations:** Individualized Drug Therapy (INDIVIDRUG) Research Program, Faculty of Medicine, University of Helsinki, Helsinki, 00290 Finland; Department of Biomedical Informatics, College of Medicine, University of Arkansas for Medical Sciences, Little Rock, AR 72205, USA; Department of Neurosurgery, College of Medicine, University of Arkansas for Medical Sciences, Little Rock, AR 72205, USA; UC Santa Cruz Genomics Institute, University of California Santa Cruz, Santa Cruz, CA 95064, USA; Department of Molecular, Cell and Developmental Biology, University of California Santa Cruz, Santa Cruz, CA 95064, USA; Wihuri Research Institute, Helsinki, Finland; Neuroscience Center, Helsinki Institute for Life Sciences, University of Helsinki, Helsinki, 00290 Finland; iCAN Digital Precision Cancer Medicine, University of Helsinki, Finland

**Keywords:** Neuro-oncology, glioblastoma, cancer relapse, genomic evolution, single-cell transcriptomics, metastasis

## Abstract

Linking glioblastoma (GBM) evolution to clinical progression is challenged by multiple factors, including tumor location for repeated sample collection, and short patient survival. In a single individual, we collected and analysed samples from 11 operations distributed across 31 months of multi-relapsing and multifocal GBM, including terminal leptomeningeal progression. All samples shared genomic ancestry of the retinoblastoma protein 1 *(RB1)* and neurofibromin 1 *(NF1)* mutations while advanced progression and extracranial metastases featured mutations of tuberous sclerosis complex 2 *(TSC2)*, *PBRM1*, *CD22* and Fanconi anemia supplementation group I (*FANCI*), correlated with clinical resistance to immunotherapies and DNA-damaging agents. Single-cell analytics revealed distinct yet reversible shifts in response to the precision medicine arsenal. GBM parenchymal dissemination and extracranial progression were associated with strengthening of neuron-like cell phenotypes. Our multidimensional study describes GBM evolution over a never reported time scale, and provides a valuable resource linking genetic, molecular, cellular and clinical progressions.

**Statement of significance:** We assembled multidimensional omics data of mutlirelapsing GBM uncovering the cascade of mutations associated with GBM progression, transcriptomic response to therapies and evolution of phenotypic cell states that ultimately lead to extracranial progression. This unique resource sheds light on GBM evolution at the genetic, transcriptomic, cellular and clinical levels.

## Introduction

Glioblastoma (GBM), one of the most aggressive forms of brain cancer, continues to present a formidable challenge to therapeutic advancement and the current standard of care regimen has not changed since 2005.^1^ Despite a staggering number of clinical trials and substantial strides in tumor biology, therapeutic drug development in GBM has remained stagnant. The neuro-oncology community has recognized the dire need for longitudinal analyses of neoplastic material throughout the treatment. This would allow to understand the aetiology of therapeutic failures and propel therapeutic advancements.^2^

Progress in single-cell omics could provide a robust surveillance platform to monitor GBM treatment response.^3^ However, all recent attempts at compiling comprehensive and exhaustive molecular profiling of GBM progression limited their sampling to one recurrence.^4 5^ Moreover, multi-omic characterization of matched primary and recurrent GBM samples demonstrated significant variability between patients with genetic evolution of both neoplastic and microenvironmental cells.^6 7 8^ In absence of repeated sampling between clinical relapses, the current consensus remains that GBM cells exhibit stronger mesenchymal (MES)-like features in response to ionizing radiation.^4^

We describe an adult patient with Li Fraumeni syndrome (i.e. germline tumor protein 53 (TP53) mutation) with GBM who underwent 11 neurosurgical resections and developed extracranial metastases. The patient received standard chemoradiation as well as immunotherapy and precision medicine. In parallel to the clinical care, fresh tumor samples were submitted to long-run multi-omic characterization over the 31-month time. Leveraging bulk and single-cell analytics, we identified the genetic, molecular, and cellular dimensions of GBM progression. Early mutational events on NF1 and RF1 were associated with mesenchymal cell state rigidity, with gradual cellular plasticity and differentiation acquired over time. We found that the two main drivers of phenotypic cell state heterogeneity were correlated with therapies and intracranial dissemination. Phenotypic cell states and therapy adaptation was mostly associated with swift enrichment of astroglial-like states. However, metastatic relapses throughout the whole brain, and extracranial ones, happened in conjunction with increased pro-neural tumor cell characteristics.

By delineating the molecular trajectories of GBM across various therapeutic regimens, we provide a roadmap for understanding the intricacies of treatment response and failure. This case report prompts a re-evaluation of our therapeutic paradigms and underscores the imperative of serial tissue sampling in deciphering the elusive mechanisms that underlie treatment resistance in GBM.

## Results

### Case report

A 25-30 year-old male with history of childhood colon cancer presented with headaches and magnetic resonance imaging demonstrated a heterogeneously enhancing 5.4 cm mass in the left temporal lobe, we identified as tumor site *A* (Figure 1A). He underwent a gross total resection, and pathology was consistent with *IDH*wt GBM. He received standard adjuvant chemoradiation but developed local and distal recurrence within 3 months of his index surgery at tumor site *B* (Figure 1A). The two recurrent lesions were resected, and cerebrospinal fluid sampling was positive for malignant cells. Patient underwent placement of an Ommaya reservoir and intrathecal therapy with the antimetabolite cytarabine as well as adjuvant radiation to the sites of recurrence. 1-year following his index surgery, he had 2 lesions concerning for distal recurrence as well as local recurrence, respectively tumor sites *C & D* (Figure 1A-C). Both sites were surgically resected with pathology consistent with recurrence for the distal site and radiation necrosis for the local site. He was then placed on the immunotherapy nivolumab but had recurrence at a new distal brain area within 1 month. He was started on a targeted therapy (palbociclib) but had rapid disease progression. Subsequently, he was enrolled in a functional precision medicine trial for GBM and was started on Osimertinib. Treatment failure occurred again, and bespoke combination therapy of Everolimus and Jakafi was initiated based on molecular and functional data.^9^ The patient also had adjuvant radiation to the multifocal site *A* where resection occurred. Eventually, treatment failure occurred again, and the tumor migrated through the cribriform plate and was removed from the nasal cavity endoscopically (site *G*) together with another intracranial recurrence distal to the index tumor (site *F*). He underwent further radiation and was eventually started on bevacizumab. On the 32^nd^ month, the patient developed a painful neck lymph node which was biopsied and confirmed recurrent extracranial GBM (site *H*). One month later, nearly 3 years after diagnosis, the patient succumbed to his disease (Figure 1B).

**Figure 1:**
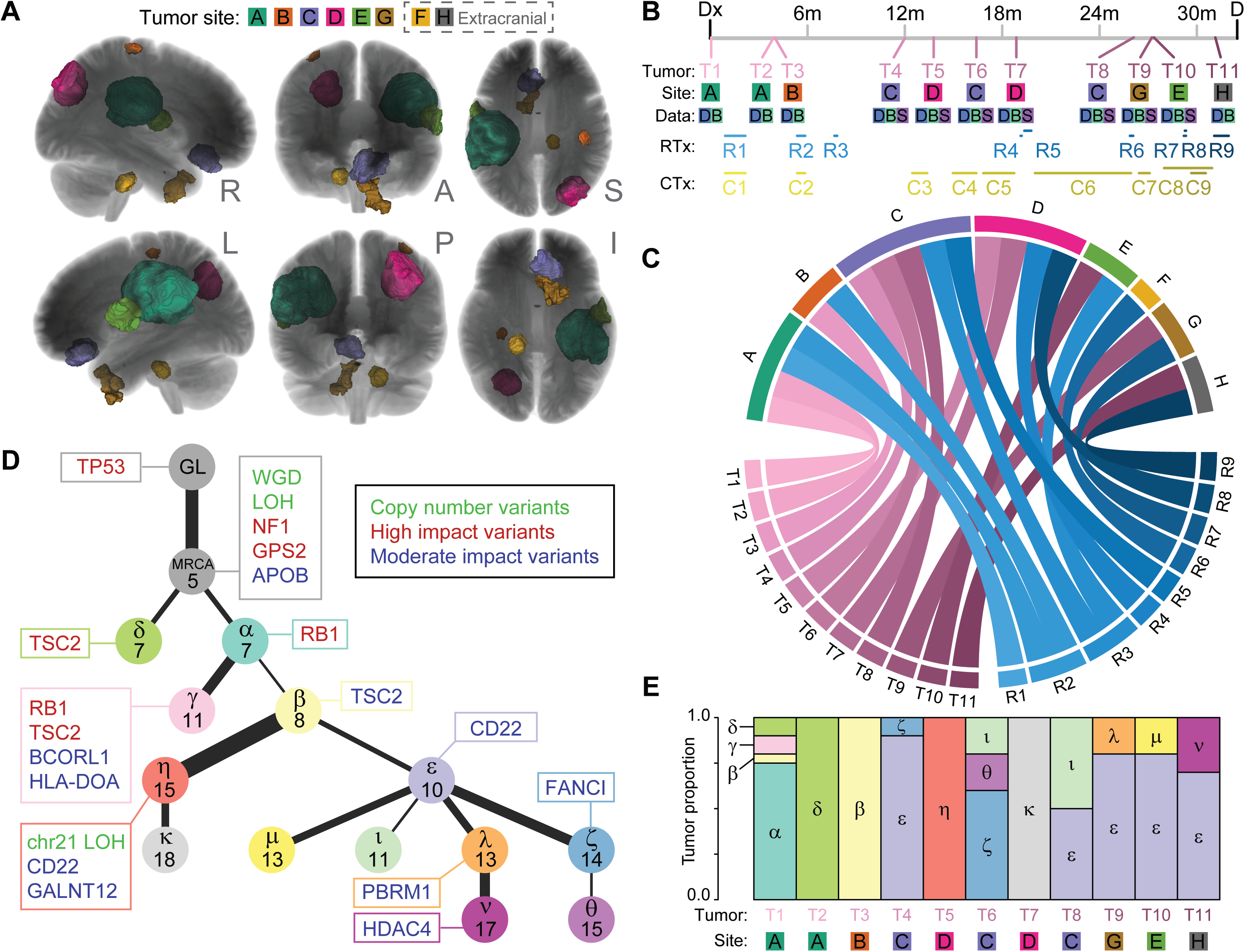
Progression of GBM with longitudinal metagenomics analysis of 11 distinct anatomical samples during the treatment continuum in an adult patient. **A**, Composite multi-planar views of the brain summarizing progression in 7 independent anatomical sites (A-G), excluding terminal extracranial progression into the lymph node (site H). Segmentations were generated from magnetic resonance scans representing the tumors at their largest size before surgical resection. R=right, L=left, A=anterior, P=posterior, S=superior, I=inferior. Tumor sites colors match those in the timeline (B) and chord plot (C). **B**, Timeline of tumor sampling and treatment from diagnosis (Dx) to death (D). Tumor samples (T1 to T11) were taken during surgical resection. The site row indicates the location of the tumors shown in panel (A). The data row shows the sequencing data produced; D=DNA sequencing, B=bulk RNA sequencing, S=single cell RNA sequencing. RTx and CTx show radiotherapy and chemotherapy windows, respectively. **C**, Chord plot linking the 8 tumors (A to H) to the samples taken (T1 to T11) and the radiotherapy received (R1 to R9). The tumor site colors match those in part A and the tumor and radiotherapy colors match those in part B. The chords are colored and ordered chronologically within their classes, with later (darker) events shown on top of earlier (lighter) events. **D**, Evolution of tumor clones reconstructed from the DNA sequencing data. Germline (GL) cells contained a *TP53* mutation, and 5 events were in all tumor cells indicating that they belong to the most recent common ancestor (MRCA). Each subsequent node is labelled with a Greek letter (α to ν) with the number of new events identified which is proportional to the thickness of the edge linking them. Notable events are labelled in matching-colored boxes. Copy number events are green, high and moderate impact variants are shown as red or blue gene symbols respectively. WGD=whole genome duplication, LOH=loss of heterozygosity. **E**, Proportion of tumor clones identified in samples T1-T11. Clone labels and colors matched those in part D and tumor and site labels match those in part B. Most samples were polyclonal, with clone ε emerging early and dominating later samples.

### Genomic landscape of GBM progression

Alongside the inherited germline *TP53* mutation, all neoplastic lesions shared only 5 somatic lesions, suggesting that they were early occurrences in tumorigenesis, although the order of events could not be determined. These lesions included severe copy number abnormalities with a median ploidy across all samples of 2.4. Tumors also exhibited widespread loss of heterozygosity (LOH) across 60% of the genome^10^ in addition to specific mutations affecting *NF1*, also co-mutated with *TP53* in cancers^11^, *GPS2* mutations lifting its tumor suppressor activity^12^ and mutated *APOB*, a biomarker of poor prognosis in many neoplasms.^13^

We detected 363 unique SNVs and 93 unique indels, with 283/363 SNVs (78%) and 15/93 indels (16%) being private to a single sample. The variant allele fraction (VAF) of private mutations was significantly lower than non-private mutations (p=8.559e-12, Wilcoxon test), suggesting that they were sub-clonal. A high-confidence subset of 36 SNVs and 6 indels was used to infer a parsimonious model of tumor evolution with 13 clones (Figure 1D). Notably, all high-impact mutations (frameshift indels and nonsense SNVs) were found in the first sample (T1), with moderate impact mutations (missense SNVs) gained throughout tumor evolution.

Interestingly, several genes were mutated in independent clones, suggesting parallel evolution. All detected clones contained at least one mutation in *RB1* or *TSC2*, suggesting that these mutations conferred a fitness advantage. Discrete inactivating *RB1* mutations were found in clones α and γ. Three different *TSC2* mutations were found in clones β, γ, and δ. Finally, two different nonsynonymous mutations in *CD22* were found in clones ε and η. Clones appeared to freely distribute throughout the brain. However, tumors in the right occipital lobe (sites *B* and *D*, collected at T3, T5, T7) were composed solely of clones β, η, and κ. Clone ε appeared relatively persistent and long-lived; first detected as the major clone in T4 (site *C*) and the major clone in the last four samples T8-T11 taken up to 18 months later (sites *C*, *G* and *E*). A unique branch in the tumor evolution emerged in sample T5 (resected from site *D*) which contained histopathological evidence of radiation-induced necrosis. The same site required re-operation (sample T7) and this relapse contained tumor cell clones of the T5 ancestry. During the last year of disease progression (T7 onwards), all resected tumors were the direct genetic evolution of the original sample T1. Remarkably, this genomic evolution included the extracranial metastases from the nasal cavity (T9, site *F*) and neck lymph node (T11, site *H*).

Altogether, genomic analyses confirmed the neoplastic evolution of 13 clones distributed across all 11 tumor samples, despite very widespread, distal brain and extracranial progression.

### Transcriptional landscape of GBM progression

To better understand the malignant progression, we next explored gene expression dynamics, employing bulk and single-cell (sc-) RNA sequencing. Bulk RNAseq data was acquired for all 11 samples (T1-11), and single cell transcriptomes from samples T4 to T10 (sites C-H, Figure 1A-C). We first evaluated the gene expression enrichment of validated GBM molecular signatures^14 15^ and phenotypic cell states.^16^ Gliomal and stromal cell proportions in bulk RNAseq were inferred with *GBMDeconvolutR*^17^ (Figure 2A). We identified a consistent trend in downregulation of GBM astro/oligodendroglial/classical gene expression signature and increasing pro-neural features over time. Meanwhile, MES-like gene expression profiles appeared upregulated at T2, T4 and T7 (Figure 2A). Interestingly, increased MES-like scores at T2 and T4 were detected in tandem with marked myeloid cell gene expression enrichment by *GBMDeconvolutR* (Figure 2B). This suggested that MES-like gene upregulation could be attributed to immunoreactivity rather than a direct readout of the tumor phenotypic cell state, as previously described.^15 18^ In samples from more advanced stages, downregulation of immune system cell markers was associated with increased proneural signatures (Figure 2A-B). This observation was in line with recent reports showing that GBM neuronal reprogramming and increased brain connectivity are fundamental drivers for intratumoral immunosuppression.^19^ Total RNAseq data was also screened for main transcriptomic cues identified in the genomic data analyses (Figure 1). For instance, gradual *NF1* gene downregulation and increased mutated *TP53* expression both matched genomic fingerprints (Figure 2C). Over the course of the tumor progression, *RB1* genetic inactivation was accompanied with detectable transcriptomic downregulation. Similarly, increased detection of *TSC2* mutation in genomics data matched the increased *TSC2* gene expression levels in bulk RNAseq samples (Figure 2C). We next verified whether transcriptomic data also exhibited evidence of response to precision medicine (Figure 2D). For instance, epidermal growth factor receptor (*EGFR*) upregulation is a commonly described therapy resistance mechanism in central and peripheral neoplasms.^20^ In our study, *EGFR* inhibition with Osimertinib (*Tagrisso*) increased *EGFR* gene expression as detected in samples collected before and after the treatment round (T7-T8) (Figure 2D). Moreover, classical/astroglial cell states are the main carriers of *EGFR* overexpression.^14 21^ We detected MES-like cell signature score decrease along AC-like cell signature enrichment following Osimertinib treatment (Figure 2D). Blockade of janus kinases 1 and 2 (*JAK1/2*) with Ruxolitinib (*Jakafi*) (at T9-11) also induced a detectable therapeutic response, with reduced gene expression of *JAK1/2* and main downstream effector *STAT3* compared to matching samples collected at earlier timepoints (T1-4) (Figure 2D).

**Figure 2:**
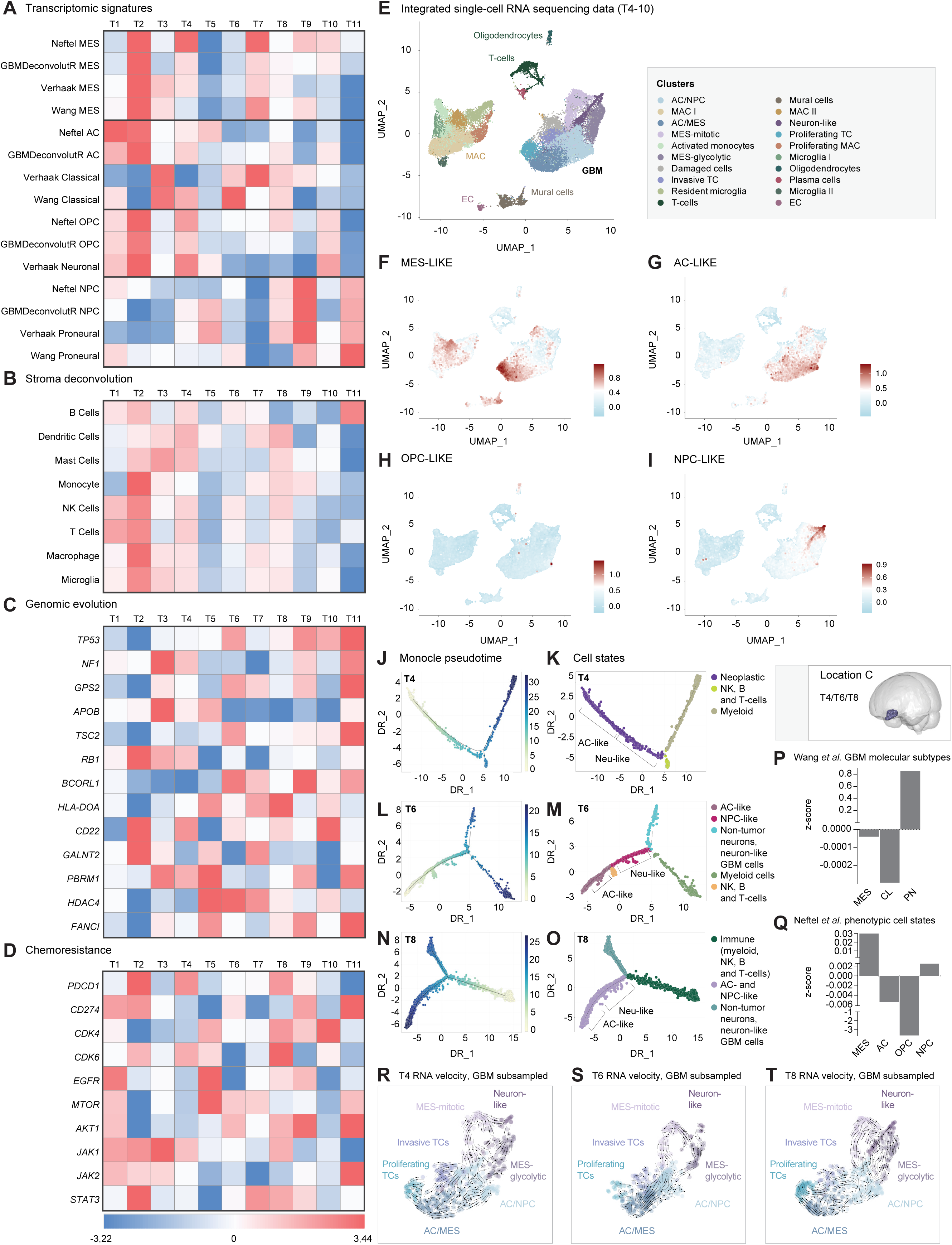
Congruence of total and single cell transcriptomic profiling on integrated cellular dynamics. **A-D**, Gene expression profiling based on bulk RNAseq data of 11 samples collected longitudinally over 31 months. All values are normalized z-scores (from blue to red) calculated from log2 gene expression levels across T1-11 samples. **A**, Cellular and molecular GBM markers were determined using published gene expression signatures e.g., phenotypic cell states, GBM molecular signature and *GBMDeconvolutR* method. **B**, Stromal deconvolutions for immune system and myeloid cells were determined using *GBMDeconvolutR*. **C**, Gene expression profiles for the genomic evolution markers identified in Figure 1. **D**, Chemotherapy response as target gene expression value. **E**, Uniform Manifold Approximation and Projection (UMAP) of the individual 41572 cell transcriptomes originating from 7 scRNAseq samples (T4-10) clustered with *Seurat*. **F-I**, GBM phenotypic cell states gene expression scores across the integrated data, including MES-like (F), AC-like (G), OPC-like (H) and NPC-like (I) GBM cells. **J-O**, *Monocle* pseudotime analyses (J, L, N) from tumor site C for biopsies 4 (T4, month 13, J-K), 6 (T6, month 17, L-M) and 8 (T8, month 27, N-O). Neoplastic and stromal cells (K, M, O) are sorted and ordered based on undifferentiated (yellow) to differentiated (blue) transcriptomic states. **H**, Composite zeta-scores of GBM molecular signatures across all bulk RNAseq data (T1-11) indicating MES cell phenotypic stability, depletion of classical/astrocytic populations and enrichment of proneural GBM signatures. **I**, Composite zeta-scores of GBM phenotypic cell states across all available scRNAseq data (T4-10), indicating robust MES cell state abundance, decreasing AC and OPC representation and increasing NPC-like over time. **J**, RNA velocity plot computed with *ScVelo* for phenotypic cell state dynamics in the tumor site C, at month 13 (T4), 17 (T6) and 27 (T8). Earlier stages of the progression are consistent with MES phenotypic stability, with brief AC adaptive response (N) and global shift towards NPC states as the tumor eventually progressed extracranially through the cribriform plate.

To increase the resolution of our bulk RNAseq analytics, we performed scRNAseq data curation of conjoined samples originating from surgeries conducted from T4 to T10. After initial data filtering excluding cells with low quality gene expression data, over 41.000 cells were integrated, clustered and labelled based on calculated cluster markers (Figure 2E). We next performed mapping of GBM phenotypic cell states across the integrated data (Figure 2F-I). For single cell omics data exploration of integrated and individual timepoints, we developed an openly accessible Shiny app web app tool called *GBMaps* discovery (http://gbmaps.it.helsinki.fi/). In these data, we detected an enrichment of MES-like genes distributed within GBM and myeloid cell populations (Figure 2F), in accordance with deconvolution analytics completed on the bulk transcriptomic samples (Figure 2A-B). AC-like cell archetypes were globally more abundant and overlapping with other cell states (Figure 2G). This was in line with the typical description of AC-like cells as a dynamic, transitional cell state existing in the tumor core.^22^ OPC-like GBM cells had very limited presence in the tumor cell clusters (Figure 2H). NPC-like gene set enrichment appears specific to cell populations of limited transcriptomic overlap with either AC-like or MES-like cell states (Figure 2I). To better understand tumor cell dynamics during GBM progression, *Monocle* data categorization was employed to classify cells based on a *stem-like* to *differentiated* transcriptomic scale (white to blue gradients, Figure 2J-M). For this, we performed focused pseudotime analyses on 3 samples (T4, 6 and 8) collected at the same brain site *C* during over a year of tumor progression. The T4 sample contained poorly differentiated neoplastic cells infiltrated by differentiated, mature myeloid and immune system cells (Figure 2J-K). At T6, the neoplastic cell fraction showed higher cellular diversity, notably through the emergence of neuron-like GBM cells sharing increased transcriptomic similarities with non-neoplastic neurons (Figure 2L-M). In the advanced T8 stage, enrichment of differentiated AC-like and neuron-like GBM cells in the tumor was associated with myeloid cell de-differentiation (Figure 2N-O). Myeloid cells differentiation relies on direct interaction with MES/stem niches but not differentiated GBM cell types, as recently reported in *IDH*-WT patient GBM.^23^ When integrating all GBM relapses in all total-(Figure 2P) and scRNAseq samples (Figure 2Q) we observed a clear trend for the enrichment of proneural and neuron-like transcriptomic features over time. In line with this, individual cellular trajectories (predicted with *scVelo* RNA velocity analysis) pinpointed initial MES to AC shifts at the first occurrence of tumor site *C* (e.g, T4 to T6) (Figure 2R-S). As the tumor progressed (T8), RNA splicing events indicated cellular trajectories steering from MES/AC-like toward pro-neural GBM cells with invasive transcriptomic features (Figure 2T). Importantly, despite their progression in extracranial spaces, both nasal (T9) and neck lymph node (T11) relapses exhibited robust up-regulation of pro-neural signatures (Figure 2A). This illustrates the importance of proneural and neuronal GBM cell differentiation as the core program associated with proximal and distal tumor dissemination.

### Precision medicine influence on GBM cellular trajectories

To determine whether cellular identities were mandated by factors such as therapies, we expanded our study of GBM evolutionary landscape to a larger timescale and integrated precision medicine as potential milestones of GBM progression. As suggested by bulk analyses, molecular and cellular fluctuations during the 3 years progression could be matching the precision medicine regimen (Figure 2A). We integrated gene expression profiles associated to targeted proteins focused on GBM phenotypic cell state abundance and score strength. For comprehensive data representation, we developed synthetic *quadriga*-plots summarizing the 4 GBM cellular components (MES-, OPC-, NPC and AC-like), their cellular abundance and signature score strength (Figure 3A). In samples T4-10 (Figure 3B) quadrigas integrate the transcriptomic response to precision medicine for molecular targets of Nivolumab (*PDCD1*, *CD274*, Figure 3C), Palbociclib (*CDK4/6*, Figure 3D), Tagrisso (*EGFR*, Figure 3E), Everolimus (*mTOR, AKT1*, Figure 3F) and Jakafi (*JAK1/2, STAT3*, Figure 3G). During the 15 months of progression captured in the scRNAseq data, AC- and MES-like states strength fluctuated the most, undergoing cell population decrease happening at T5, T6, T7 and T8, matching precision medicine protocols applied to the patient (Figure 3B-G). Interestingly, NPC-like archetypes were seemingly unaffected by chemotherapies. AC-like cell populations increased in number and featured increased expression of precision medicine targets compared to other subtypes (Figure 3C-G). For instance, the tumor response to immune checkpoint inhibitors was clearly detected in MES-like cells. Treatment with Nivolumab downregulated *CD274* in MES archetypes, which lasted months after the treatment round (Figure 3C). Unfortunately, MES cells eventually recovered CD274 expression as detected in sample T8 onwards, potentially matching the immune system cell exhaustion detected in bulk RNAseq samples (Figure 2A). Next, targeting EGFR with Tagrisso (between T6 and T7) induced a detectable molecular and cellular response (Figure 3E). In accordance with bulk RNAseq data and previous reports, MES-like cell population decreased in favor of more abundant AC-like cell states.^14,16^ These AC-like “survivors” upregulated *EGFR*, possibly as a compensation mechanism for the receptor inhibition (Figure 3E).^20^ Similarly, the response to the mTOR inhibitor (Everolimus, Figure 3F) and Janus kinase (Jakafi, Figure 3G) mostly affected MES-like cell populations. This led to transient decreased genes expression (*MTOR*, *AKT1 JAK1/2* and *STAT3*) in this MES-like cells. Both Everolimus and Jakafi induced a transient adaptive cellular compensation with increased AC-like numbers and gene signature score. Altogether, those cellular and molecular dynamics correlated with precision medicine supported preferential cellular adaptations occur in response to treatment. Tumor dissemination however appears to be linked with increased neuronal transcriptomic mimicry, which aligns with preclinical studies linking GBM neuronal connectivity driving tumor invasion.^24 25 26 27^

**Figure 3:**
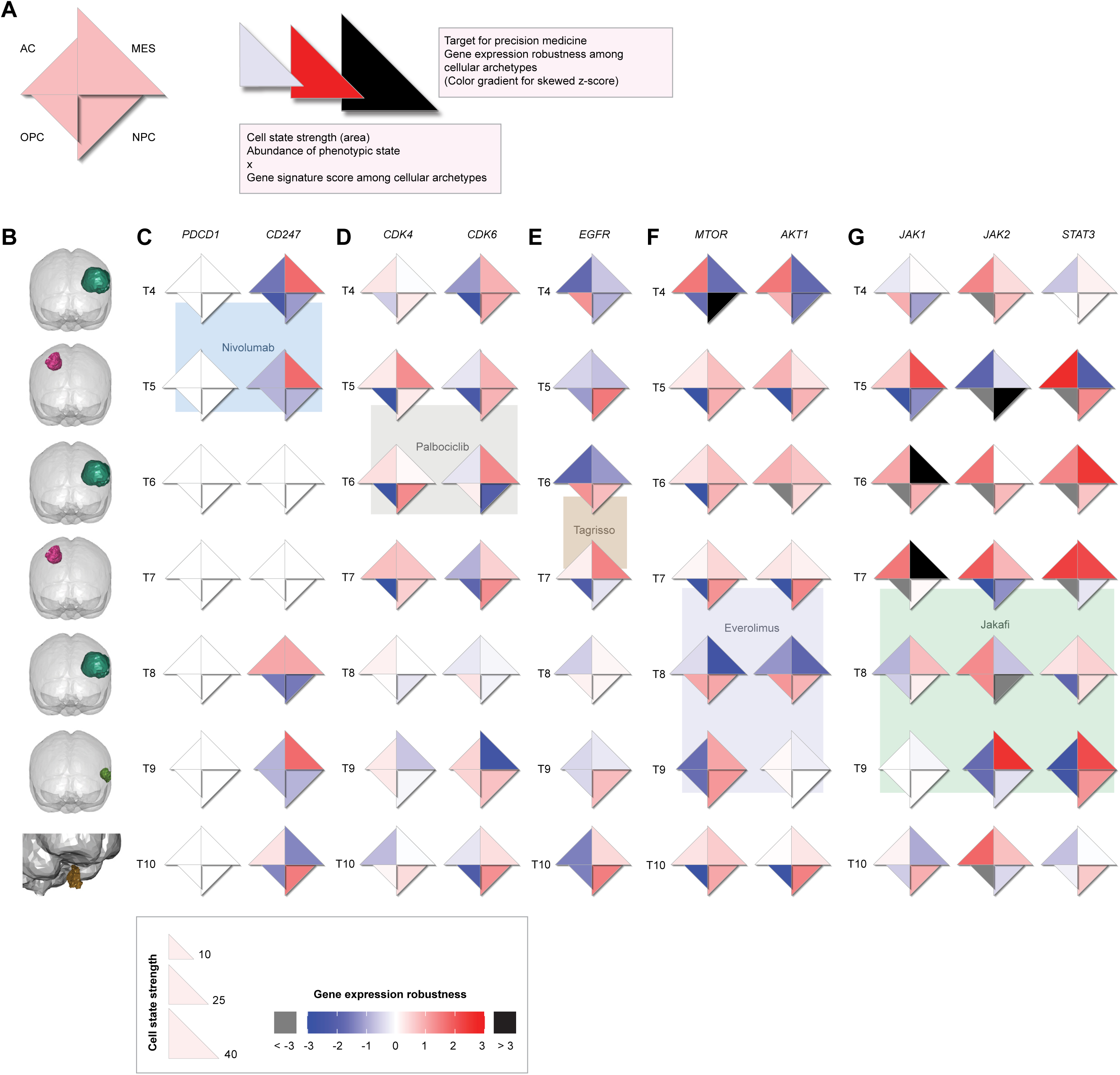
GBM evolution follows therapy response dynamics. **A**, Summary of the *quadriga* plots representing MES, AC, OPC and NPC phenotypic cell states and associated therapy target gene expression. Cell state strength in a sample integrates abundance of cell archetypes and gene expression signature (z-score) per cell, represented by quadrants of various areas calculated according to the state strength. Precision medicine target genes expression are expressed as skewed z-scores integrating positive cell numbers and gene expression levels per phenotypic cell state. Values are expressed gradient of blue (-3 to 0), white (no change) and red (0 to 3). Extreme negative (<-3) and positive (>3) values are plotted in grey and black, respectively. **B**, Representation of the operated brain areas for samples T4-10. **C**, *quadriga* plots for the cellular and transcriptomic response to Nivolumab (*PDCD1, CD274*) in samples T4-10. **D**, *quadriga* plots for the cellular and transcriptomic response to Palbociclib (*CDK4, CDK6*) in samples T4-10. **E**, *quadriga* plots for the cellular and transcriptomic response to Tagrisso (*EGFR*) in samples T4-10. **F**, *quadriga* plots for the cellular and transcriptomic response to Everolimus (*MTOR, AKT1*) in samples T4-10. **G**, *quadriga* plots for the cellular and transcriptomic response to Jakafi (*JAK1, JAK2, STAT3*) in samples T4-10.

## Discussion

Experimental data in cells and preclinical models have uncovered the molecular dynamics regulating phenotypic states of GBM stem cells during progression. Although providing a new dimension to our understanding of intra- and inter patient variability, validation in longitudinally sampled human material is challenged by the short survival. Our multi-omics study encompassing three years of GBM progression in a patient is among the firsts addressing this gap. Interestingly, we found that although emerging from distinct anatomical locations compared to the primary tumor site, all neoplastic lesions were characterized by the same primordial genetic signature. As previously described for Li-Fraumeni patients^28^, mutations affecting *TP53* and *TSC2* genes were detected in all tumors. During the next step of the tumor progression, *NF1* and *RB1* gene mutations appeared following the typical description of the cell of origin for GBM.^29^ Interestingly, late-stage emerging mutation on *FANCI* has been recently suggested to be correlated with advanced GBM progression.^30^ This prompted us to interrogate bulk and single-cell transcriptomics data to verify the molecular and cellular dynamics during progression. First, independent analyses of bulk and single-cell transcriptomes led to matching conclusions, especially after implementing gene expression deconvolution on bulk data. This suggests that despite the significantly higher resolution of single-cell omics, the benefit is not matching the costs (infrastructure, financial, and time) for clinical diagnosis. Beyond this new technical insight, we demonstrate the great molecular and cellular adaptability of GBM, when disseminating in the brain and in response to therapies. For instance, EGFR precision medicine impacted astrocyte-like GBM cell populations the most, as their growth is highly dependent on EGFR activation.^31^ In turn, we observed a rapid tipping towards MES-like cell archetypes, whose growth rely on alternative pathways such as JAK/STAT.^32^ As a clinical response, we detected that therapies such as everolimus, targeting mTOR/JAK/STAT signalling showed excellent response in reducing MES-like cell populations and silencing JAK/STAT amplification. However, tumor cell adaptability appeared to consistently kick-in, supporting the generation of chemoresistant clones. Eventually, we confirmed independent experimental studies suggesting that enrichment of neuronal and proneural features supported GBM metastasis in the central nervous system. In addition, we show for the first time that extracranial tumors also feature neuronal archetypes, suggesting that this distinct cell state fuels intra- and extracranial metastasis.

## Methods

### Patient consent and ethical permits

The study was conducted according to the guidelines of the Declaration of Helsinki and approved by the Institutional Review Board of University of Arkansas for Medical Sciences (IRB #228443 approved on 13 November 2018). The use of human brain tissue was coordinated by the University of Arkansas for Medical Sciences (UAMS) Tissue Biorepository and Procurement Service (TBAPS) for biospecimen banking following the ethical and technical guidelines on the use of human samples for biomedical research purposes under the Institutional Review Board-approved protocol (IRB #228443) and general consent form. Patient tissues were collected at the University of Arkansas for Medical Sciences after informed patient consent was obtained, and all samples were de-identified before processing.

### Tissue collection, sample preparation and sequencing Single-cell transcriptomics

Fresh surgically resected tumor tissue was placed in sterile phosphate-buffered saline and taken immediately to the lab after TBAPS de-identification process. The tissue was distributed and kept at 4 °C. Immediately after tumor reception, the specimen is washed with phosphate-buffered saline (PBS) to remove excess of blood and dissected to remove any visible necrosis.

Tumor cut in pieces (<1 cm) and digested with collagenase-based buffer, overnight at 4 °C, on an orbital shaker (>20 rpm). The digestion buffer contained type II and IV collagenases (0.18% wt/vol each, # LS004176, Worthington and #17104-019, Gibco respectively), DNAse (0.36% wt/vol, #DN25-1G, Sigma), DMEM (50 mL, #10-013-CV, Corning), penicillin/streptomycin (5 mL, #15140-122, Gibco). The sample was then incubated for 45’ at 37°C in water bath under agitation. The resulting cell suspension was passed through 70 µm cell strainer (#22-363-548, Fisher Scientific) and washed with either Hank’s Balanced Salt Solution (HBSS) or Ca^2+/^Mg^2+^-free D-PBS (3 mL). Ficoll-Paque (3 mL, #17-5446-02, GE Healthcare) was used to layer the cell suspension into 15 mL conical tubes. The tubes were centrifuged at 300 x g for 35’ (Sorvall ST 16, ThermoFisher) at 18-20 °C in a swinging bucket rotor. The upper layer was discarded, leaving the interface/cell layer undisturbed. The entire interface was transferred with a minimal amount of Ficoll-Paque to a new 15 mL conical tube. At least 3 volumes (6 mL) of D-PBS were added to each tube. The cells were suspended by gently pipetting them up and down and the tubes were centrifuged at 100 x g for 10’ at 18-20 °C. The pellets were resuspended in HBSS (6 mL), and centrifuged at 100 x g for 10’ at 18-20 °C. The supernatant was removed and discarded. The cells pellet was resuspended in a small volume of D-PBS and counted using the AO/PI primary cells settings from DeNovix CellDrop (CD-AO-PI-1.5, DeNovix Inc.). Cells were washed in D-PBS to remove debris until viability reached 70% or higher. Cells were then centrifuged at 250 x g for 10’ and re-suspended in D-PBS supplemented with 0.04% (w/v) bovine serum albumine. To determine cell concentration and viability, cells were stained with ReadyProbes™ Cell Viability Imaging Kit, Blue/Green (# R37609, Thermo Fisher Scientific), and manually counted using hemocytometer on an EVOS M7000 microscope (Thermo Fisher Scientific).

Immediately following cell counting, samples were processed using Chromium Next GEM single cell 3_’ Reagent Kits v3.1 (Dual Index) as described in the manufacturer ‘s instructions. In brief, aiming for 8000 cells per library, single cell suspensions with more than 70% live cells were loaded onto Chromium Controller (10X Genomics) to generate gel beads-in-emulsions (GEMs). Then co-partitioned cells were lysed, primers were released from Gel Beads, and barcoded full-lengh cDNA was produced and amplified. 3’ gene expression libraries were generated from cDNA by fragmentation, end repair, A-tailing, adaptor ligation and index PCR amplification. The concentration and size distribution of final libraries were assessed by Qubit™ _1X dsDNA HS Assay (# Q33231, Thermo Fisher Scientific) and the Fragment Analyzer System (Agilent). Libraries were sequenced either on a NextSeq500 or NovaSeq 6000 (Illumina) with paired-end mode (read1: 28 cycles, read 2: 90 cycles, i7: 10 cycles, i5: 10 cycles) to generate a minimum of 20,000 read pairs per cell.

### Tumor preparation for bulk RNA sequencing

Tumor samples preserved in formalin fixed paraffin were processed and sequenced by Tempus (Chicago, IL, USA). Tumor DNA was extracted from tumor tissue sections using proteinase K digestion. Only sample containing tumor cellularity higher than 20% was next processed for sequencing. Chemagic360 instrument was utilized to carry out the extraction of total nucleic acid through a magnetic bead purification protocol specific to the source. The nucleic acid was quantified by using Quant-iT Ribogreen RNA Kit (Life Technologies, Carlsband, CA, USA). Quality was confirmed using a LabChip RNA High HT Pico Sensitivity Reagent Kit (PerkinElmer, Akron, OH, USA). For the library construction, 100 ng of RNA per tumor sample was fragmented with heat in the presence of magnesium to an average size of 200 base pairs. The RNA then underwent first strand cDNA synthesis using random primers, followed by combined second strand synthesis and A-tailing, adapter ligation, bead-based cleanup, and library amplification. After library preparation, samples were hybridized with the IDT xGEN Exome Research Panel. Target recovery was performed using Streptavidin-coated beads, followed by amplification using the KAPA HiFi Library Amplification Kit. The RNA libraries were sequenced to obtain approximately 65 million reads on an Illumina HiSeq 4000 System.

### Somatic variant calling pipelines

Single nucleotide variants (SNVs) were called using *Strelka2*^33^ and filtered using *FiNGS*^34^ using default parameters with a maximum depth of 10000 reads and minimum variant allele frequency (VAF) of 0.01. Short insertions and deletions (indels) were called by taking the union of the results of *Freebayes* and *Pindel*.^35^ Indels were manually filtered after reviewing all potential sites in IGV.^36^ To recover subclonal variants, we extracted read counts from the site of every called variant in every sample and filtered using these criteria; 30x or deeper coverage in at least 10/11 samples, a minimum VAF of 0.05 and a minimum of 10 supporting reads in at least one sample. This removed variants that were not informative for tumor evolution as they were either consistent false positives or subclonal in every sample.

### Copy number, LOH, and Purity

*ASCAT*^37^ was used to calculate copy number, loss of heterozygosity (LOH), and the proportion of tumor cells (purity). Purity estimates were compared to the VAFs of homozygous SNVs in regions of LOH that were clonal in all samples. The median sample purity was 0.6 (range 0.1 to 0.8). Purity-corrected VAFs were produced by dividing the raw VAF value by the purity of that sample.

### Phylogenetics and tumor evolution

Hierarchical clustering was applied to the purity corrected VAFs of filtered variants. Variants were assigned to clones based on the chronological order they appeared to create a parsimonious phylogeny using the minimal number of clones. The proportion of each clone in a sample was estimated using the purity-corrected VAFs and adjusting for any LOH and copy number affecting each variant.

### Composite Z-score calculations

To determine GBM molecular^15^ and phenotypic cell states^16^ characteristics, we calculated composite evolutionary z-scores (z_evo_) using the following formula:

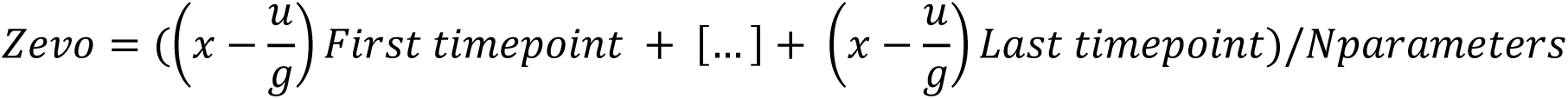

With x = molecular/cell signature score, u = mean gene expression value per bulk sample/cell state, g = standard derivation per bulk sample/cell state.

### Quadriga plots

To calculate the cell state strength per quadrant, median log2 gene expression for all genes of phenotypic cell state was calculated in all neoplastic cells, per timepoint. Summed phenotypic cell state scores for cells with a score > 0 (including cells with hybrid/dual states in the calculation) was used to normalize (%) the relative abundance of each state. Normalized values were used to set the quadrants’ area. For the precision medicine gene expression response (up- or downregulation), z-scores for the log2 gene expression per timepoint were calculated across all 4 cell states. These scores were used to assign the color gradient (blue to red, extreme negative and positive values in grey and black, respectively). The code to generate *quadriga* plots in R is available upon request.

### Analysis of single-cell RNA sequencing (scRNAseq) data

Raw count matrices were imported to an *R* V4.5.3-environment using the *Read10X()*-function by *Seurat* V5.4.0. The *Seurat* package was further used for preprocessing and clustering of the data. First, the data were filtered to include cells with minimum 200 features (genes) and genes encountered in at least 3 cells. Next, cells were filtered based on their number of unique RNA species and based on their mitochondrial (pt.mito) and ribosomal gene percentages (pt.rRNA). The filtering parameters were chosen in a sample-specific manner, and the sample-specific values can be found in Supplementary Table 1. With these filtering parameters, 14.0-31.1% of the cells were filtered out, and remaining cells were further normalized on a logarithmic scale of 10000. The 2000 most variable features were retrieved using the variance-stabilizing transformation algorithm.

Data integration anchors were calculated using 50 dimensions and the samples were then integrated using the *IntegrateData()*-function. The data were scaled regressing for RNA counts, pt.mito, and pt.rRNA. Next, principal component analysis was run, and clusters were calculated using 40 dimensions and a resolution of 0.6 using the *FindClusters()*-function. The resulting 20 clusters were visualized using Uniform Manifold Approximation and Projection (UMAP) and labeled based on differentially expressed genes (DEGs) retrieved using the *FindMarkers()*-function with the minimum part of expressing cells set to 0.25 (Supplementary Table 2).

### RNA velocity

RNA velocity trajectories were calculated with *scVelo* V0.2.5 in a *Python* V3 environment based on the ratio of unspliced to spliced RNA counts. Briefly, splicing ratios were calculated using *Velocyto.py* V0.17.16 with the *Puhti* supercomputer provided by the Finnish IT Center for Science (CSC, Espoo, Finland). Next, the splicing counts together with UMAP embeddings calculated in *Seurat* V5.4.0 were imported to a *Python* V3 environment and further processed to visualize velocity trajectories using *scVelo* V0.2.5 using the stochastic model.

### Analysis of bulk RNA sequencing (RNA-seq) data

Total RNAseq data were processed using the *Tempus xR* pipeline version 1 and 2. (Chicago, IL, USA). Samples T1-7 (“TL-19-4DACEA” “TL-20-003856” “TL-20-533774” “TL-20-69E5E8” “TL-20-85DE5A” “TL-21-2396CE” “TL-21-F71366”) were sequenced on the V.1 pipeline, and samples collected after June 2021 e.g., T8-11 (“TL-21-EXJKSKD4” “TL-21-WBPCZ3JG” “TL-22-GK9JIUPV” “TL-22-GVFT5YJY”) on the V.2 pipeline. Batch correction factor was applied to the transcripts per million values for each gene in samples 8-11. Data were imported to *R* V.4.2.1 for data visualization using the *tidyverse* and *ggrepel* packages.

## Data availability

Single cell transcriptomics data annotation, visualization and integration are available in the discovery app *GBMaps* (http://gbmaps.it.helsinki.fi/). All other data is available from the corresponding authors upon request

## Acknowledgements

We are extremely grateful to the patient and his family who participated in this research and made this work possible. We acknowledge the support of the University of Arkansas for Medical Sciences, which provided laboratory facilities for this study, the CSC IT Center for Science (Finland) for providing the supercomputer Puhti services, and University of Helsinki IT Services for the Linux server administration of *GBMaps*.

This study was supported by funding from the University of Arkansas for Medical Sciences, the Research Council of Finland, the Finnish Cancer, Magnus Ehrnrooth and K. Albin Johansson foundations. V.L.J. acknowledges the support of the Lundbeck Foundation. M.H.L. acknowledges the financial support from the Nylands Nation, the Doctoral Programme in Integrative Life Science (University of Helsinki), Biomedicum Helsinki Foundation, Orion Research Foundation sr, and Finska Läkaresällskapet.

